# Variation in Aerosol Production Across Oxygen Delivery Devices in Spontaneously Breathing Human Subjects

**DOI:** 10.1101/2020.04.15.20066688

**Authors:** Theodore J. Iwashyna, Andre Boehman, Jesse Capecelatro, Amy M. Cohn, James M. Cooke, Deena Kelly Costa, Richard M. Eakin, Hallie C. Prescott, Margaret S. Woolridge

**Affiliations:** VA Center for Clinical Management Research, Ann Arbor Veteran Affairs Health System, Ann Arbor, Michigan, USA; Medicine, at the University of Michigan, Ann Arbor, Michigan, USA; Mechanical Engineering, at the University of Michigan, Ann Arbor, Michigan, USA; Industrial & Operations Engineering, at the University of Michigan, Ann Arbor, Michigan, USA; Family Medicine, at the University of Michigan, Ann Arbor, Michigan, USA; Learning Health Sciences, at the University of Michigan, Ann Arbor, Michigan, USA; Nursing, at the University of Michigan, Ann Arbor, Michigan, USA; Respiratory Care, at the University of Michigan, Ann Arbor, Michigan, USA; Aerospace Engineering at the University of Michigan, Ann Arbor, Michigan, USA

## Abstract

We sought to assess whether HHFNC results in greater production of aerosolized particles than 6 liters per minute nasal cannula, using state-of-the-art techniques of aerosol measurement, in spontaneously breathing human volunteers in a simulated hospital room.

For each volunteer, we first measured background aerosol levels in the room immediately prior to testing. We then measured aerosol levels while the healthy volunteer laid in bed - - with the head of bed at 30 degrees - - wearing the following oxygen delivery devices: (a) 6L/min nasal canula (NC) with humidification; (b) non-re-breather mask (NRB) with 15L/min gas flow, non-humidified; (c) HHFNC with 30L/min gas flow; (d) HHFNC with 60L/min gas flow. Two scanning mobility particle sizing (SMPS) systems (TSI 3080/3030, TSI 3080/3750) were used to measure aerosols 10 to 500 nanometer (nm) in size for each of the oxygen delivery devices.

There was no variation in aerosol level within patients between room air, 6 L/min NC, 15 L/min NRB, 30 L/min HHFNC, and 60 L/min HHFNC, regardless of coughing.

---

Heated high-flow nasal cannula (HHFNC) has emerged as an important life support tool for patients with acute hypoxemic respiratory failure, especially for patients in whom intubation may be riskier or incompatible with their goals of care. It may also have an important role in facilitating extubation. ^1,2^

Despite the central importance of hypoxemic respiratory failure as a mechanism of mortality in COVID19, several authorities have recommended against the use of heated high-flow nasal cannula in patients with suspected or confirmed COVID19. ^3-5^ These recommendations are driven in part by data of dispersion and stability of SARS-CoV-2 in clinical spaces ^6-8^, and fears that high flow rates will increase aerosolization of SARS-CoV-2, including by data from an artificial simulator using a smoke dispersion model. ^9,10^ In contrast, lower flow oxygen is routinely assumed to present an acceptable risk of aerosol dispersion. Therefore, in order to inform care policy for COVID19 patients at the VA Ann Arbor and University of Michigan regarding infection control under oxygen support, we sought to assess whether HHFNC results in greater production of aerosolized particles than 6 liters per minute nasal cannula, using state-of-the-art techniques of aerosol measurement, in spontaneously breathing human volunteers in a simulated hospital room.

## Methods

This work was conducted as a quality improvement project in order to inform care policy for COVID19 patients at the VA Ann Arbor and University of Michigan regarding infection control under oxygen support, and reviewed and deemed not regulated by the University of Michigan Institutional Review Board (HUM00179622).

For each volunteer, we first measured background aerosol levels in the room immediately prior to testing. We then measured aerosol levels while the healthy volunteer laid in bed—with the head of bed at 30 degrees—wearing the following oxygen delivery devices: (a) 6L/min nasal canula (NC) with humidification; (b) non-re-breather mask (NRB) with 15L/min gas flow, non-humidified; (c) HHFNC with 30L/min gas flow; (d) HHFNC with 60L/min gas flow. Two scanning mobility particle sizing (SMPS) systems (TSI 3080/3030, TSI 3080/3750) were used to measure aerosols 10 to 500 nanometer (nm) in size for each of the oxygen delivery devices. Additional measurements were made during HHFNC use for larger particles (from 10 to 10,000 nm in size) using two particles counters (PCs; TSI 3030, TSI 3330). For reference, testing was performed with a low aerosolization control (HEPA filter) and 2 high aerosolization controls (with a candle burning, and with a water nebulizer).

The SMPS aerosol measurement probes were placed in two locations (**Figure 1**) selected for their clinical relevance: (#1) affixed to the bed rail beside the patient’s head (mimicking where someone would stand to perform suctioning or oral care). The probe at location #1 was also relocated to a second position (#2) to approximately 10 cm from the patient’s mouth for each oxygen delivery device test. The particle counters measurements were made at position (#2).

**Figure 1:**
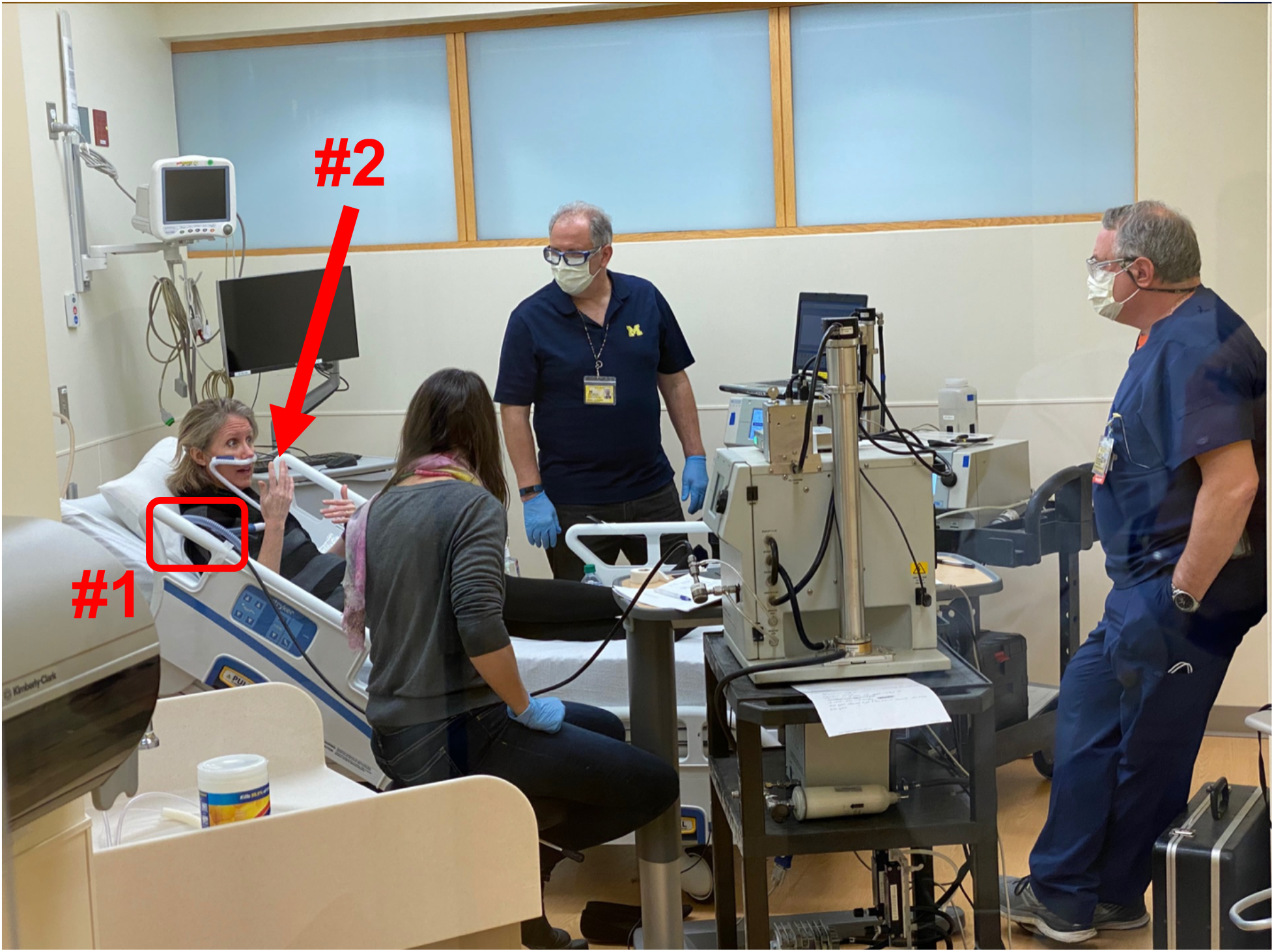
Examples of measurement location and test set-up. All photographed individuals are named authors on the manuscript and this is used with their permission.

Aerosol measurements were obtained using the SMPS and PC systems, which reliably assess the distribution of particles of size 10 to 10,000 nm, the relevant range for human coughing.^11^

Each sample was collected for three minutes and data were acquired for approximately a 10-minute period for each oxygen delivery device. The healthy volunteers were instructed to intentionally cough at each setting; spontaneous coughing occurred for some volunteers throughout and was not suppressed in any way. The simulated hospital room matched the dimensions, shape and layout of a single occupancy hospital room with all equipment, monitors and computers standard to this setting. Standard wall gas supplies and regulators were used with compressed air substituted for supplemental oxygen. Investigators wore standard surgical face masks and remained in the room and stationary for each extended sampling period to minimize air disturbance. We recorded each sampling period (video and audio) for reference and data archiving.

## Data Analysis

For each aerosol sample, the particle size distribution, the total number of particles, the total mass of particles and the average particle size were measured

## Results

Four healthy volunteers from the authorship group participated, 2 male (Patients 1 and 2) and 2 female (Patients 3 and 4). None had known lung disease, recent exposure to COVID-19 or other sick contacts, or recent travel to a known area of transmission. No testing occurred with the volunteers wearing masks.

**Figure 2** shows particle mass concentrations among the first 3 patients; note the log vertical scale. As seen in the figure, there was variation in baseline measurements (i.e., room air) day-to-day. Samples of the room air are included for reference in **Figure 2** at measurement location #1, the head of the bed. (Patients 1 and 2 were done on one day, Patient 3 on another.) As seen in the bottom center of the figure, HEPA filtration caused an order of magnitude decrease in particle concentrations, and the burning of a candle caused an order of magnitude increase in particle concentrations. Similar increases were seen with the nebulizer (data not shown). There was no variation in aerosol level within patients between room air, 6 L/min NC, 15 L/min NRB, 30 L/min HHFNC, and 60 L/min HHFNC, regardless of coughing. Consistent results are shown in **Figure 3** for the particle number concentration (via SMPS), and in **Figure 4** for average particle diameter (via SMPS).

**Figure 2:**
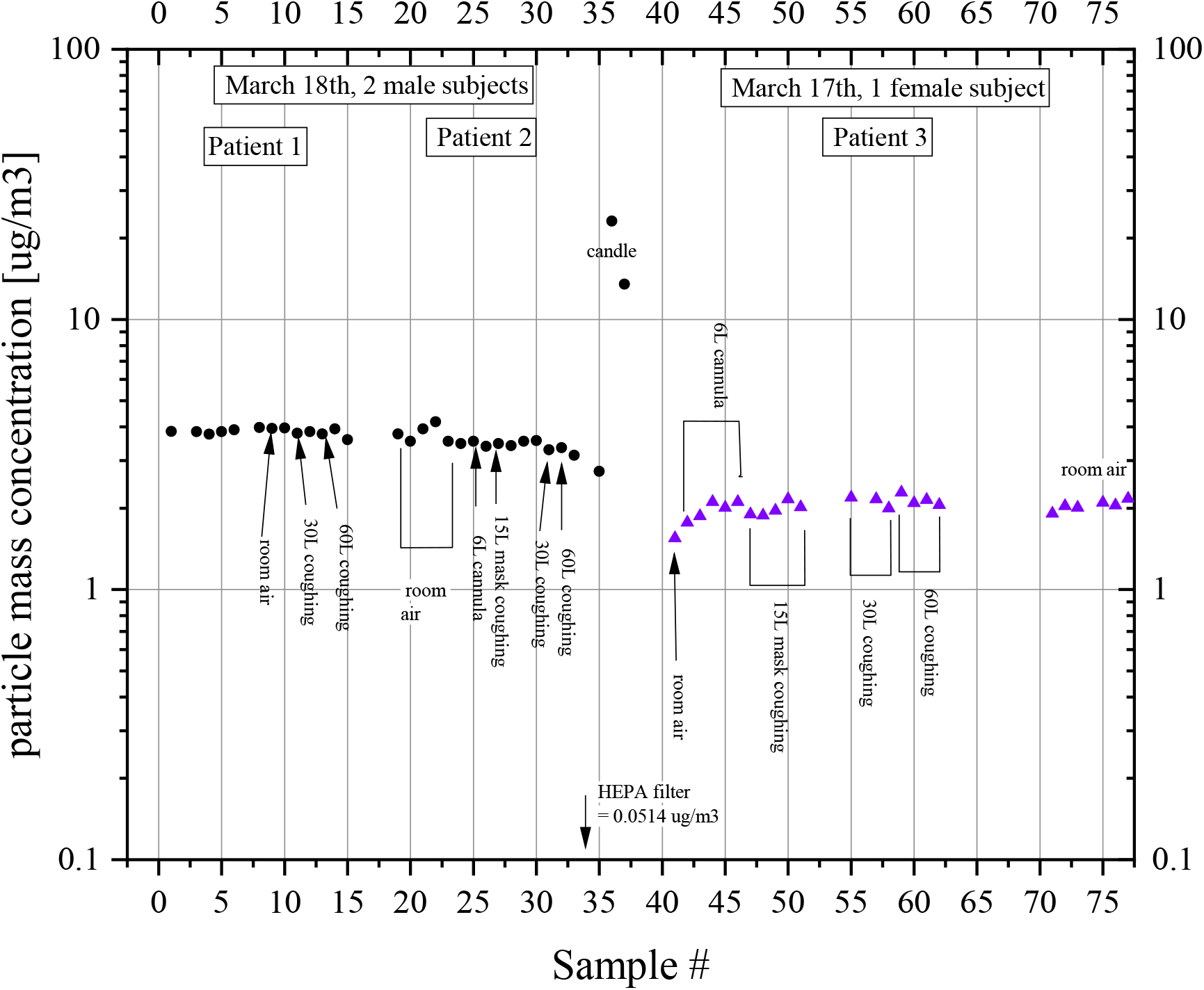
Particle mass concentration (SMPS)

**Figure 3:**
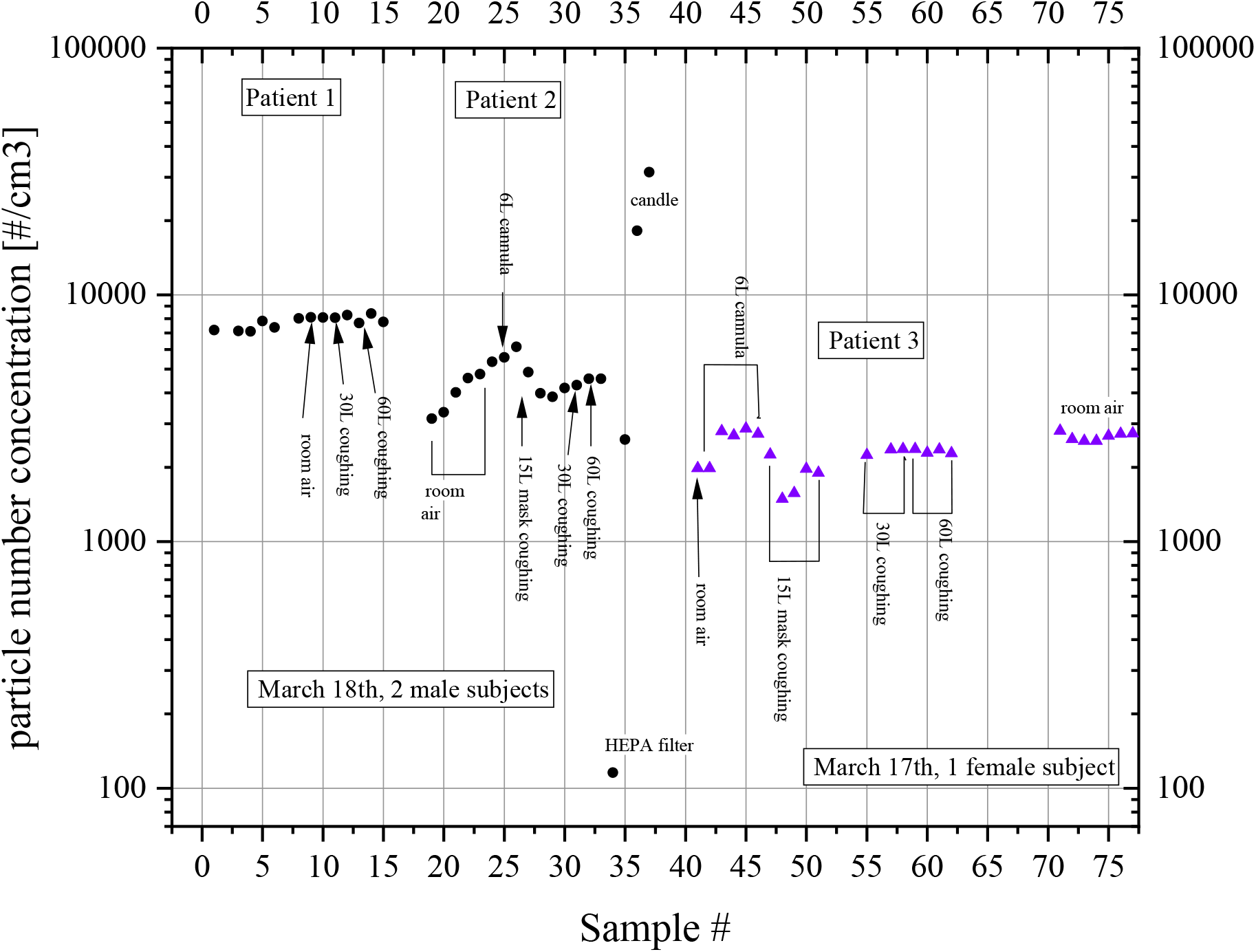
Particle number concentration (SMPS)

**Figure 4:**
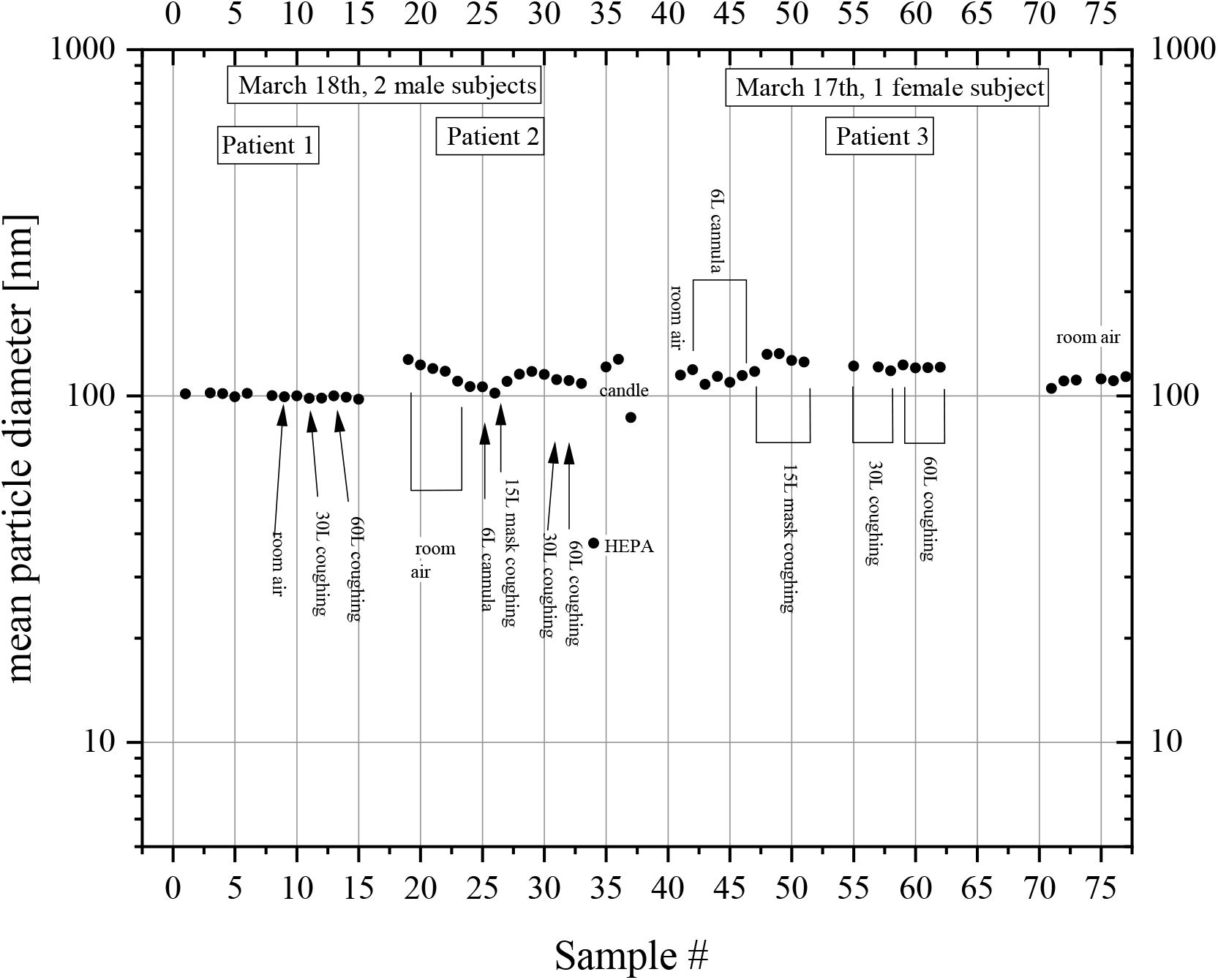
Average particle diameter (SMPS).

Measurements with the Patients 1, 3, and 4, that included broader range of particles, up to 10,000 nm (=10 micron), also showed consistently no increase above the room levels of particle number concentration during HHFNC use at 30 L/min or 60 L/min (**Appendix 1**). (These tests were repeated for Patients 1, 3, and 4 on a third testing day with the particle counters at position #2 in **Figure 1**.)

## Conclusion

In these healthy human volunteers, there was no evidence of increased aerosolization with nasal cannula, non-rebreather mask, or heated high flow nasal cannula in the particle size range of 10 nm to 10000 nm. This may contrast with the interpretation of another study that used mannequins and an intentional addition of smoke to their model of the right middle lobe of the lung as a measure, although the actual dispersion distances shown in that study are similar. ^9^ In contrast, Loh *et al* reported visible assessments of gargled food dye in each of 5 patients who coughed twice, which showed highly variable changes in coughing distance from a reduction of 2 cm to an increase fo 84 cm using HHFNC compared to spontaneous breathing.^12^ Our data are consistent with Leung *et al*’s culture-based data showing no increased dispersion in infectious organisms with the use of HHFNC in ICU patients with pneumonia.^13^

A limitation of the present study is the modest numbers of volunteers, with consequent limitations in the degree of variation in naso/oral structures. None of these human subjects had an active viral syndrome—although also neither did any of the test mannequins in prior data. These data do not speak to the aerosolization potential of other modalities, nor to debates about the relative merits of early intubation and volume limitation versus spontaneous breathing with heated high flow nasal cannula in the acute respiratory distress syndrome. Debate about the correct biophysical conceptualization of droplet spread remains. ^14^ Despite these limitations, these data doprovide direct evidence that there may be limited differences in aerosolization between alternative modes of oxygen delivery at clinically relevant distances and positions for routine care.

## Data Availability

Data will be provided to potential collaborators.

## Acknowledgements

We would like to thank Gene Kim and Lee Kamphuis for their administrative support.

## Appendix Data from Broader range Particle Counters for Patients 1, 3, and 4

**Figure A1:**
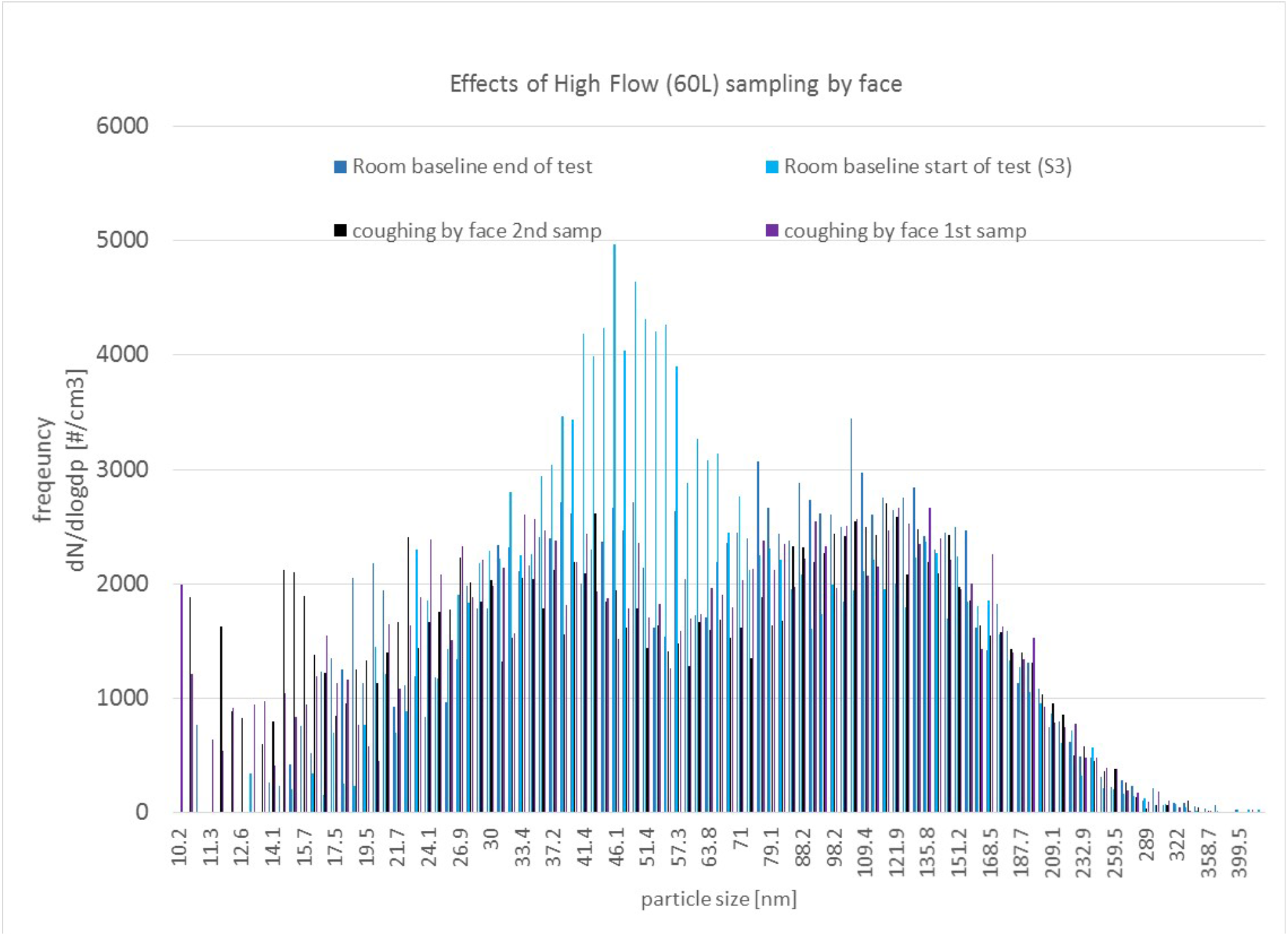
Particle size distributions (SMPS) of room air and 60 L/min HHFNC.

**Figure A2:**
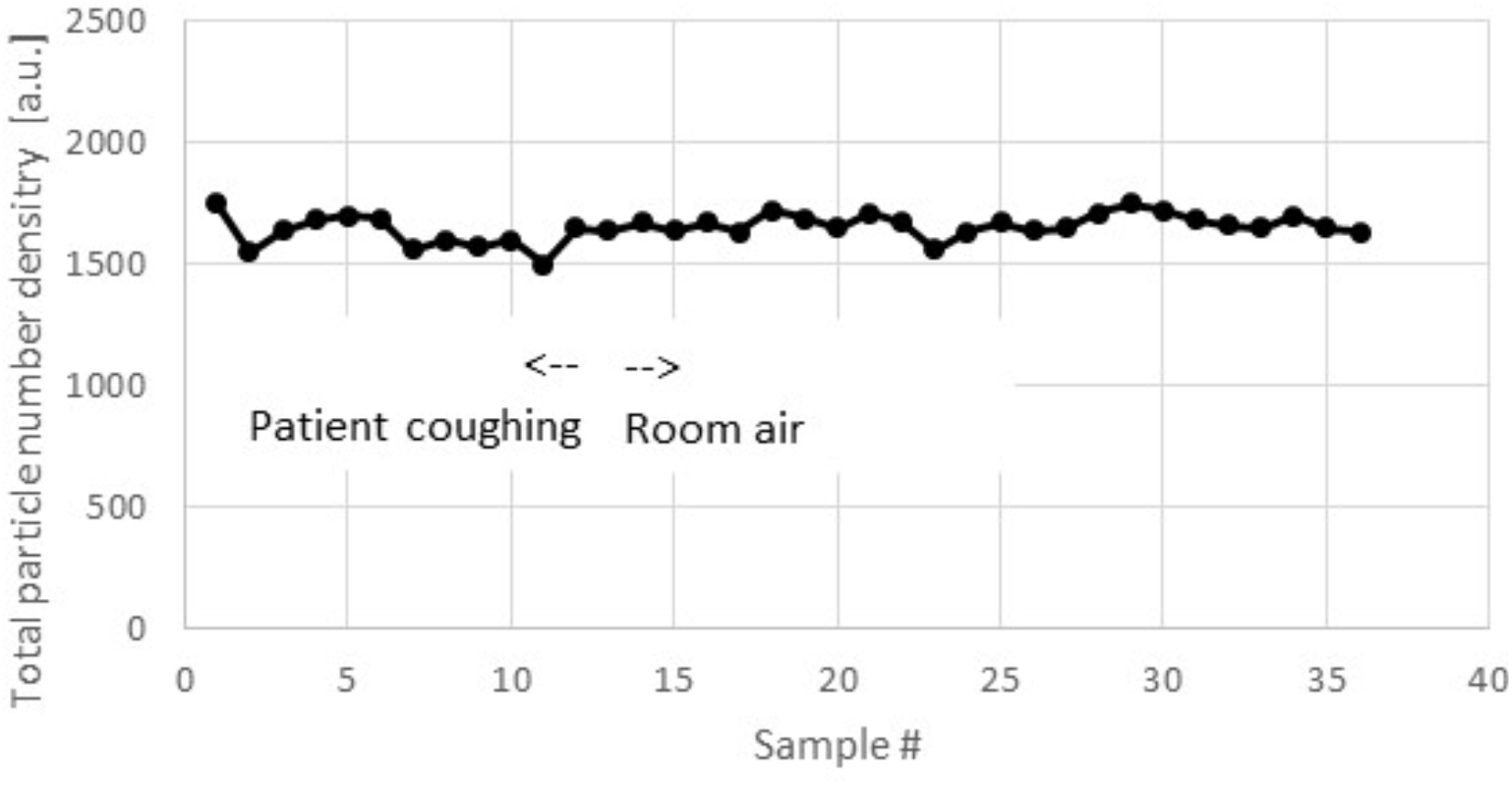
Particle number concentration (PC) room air and 60 L/min HHFNC.

**Figure A3:**
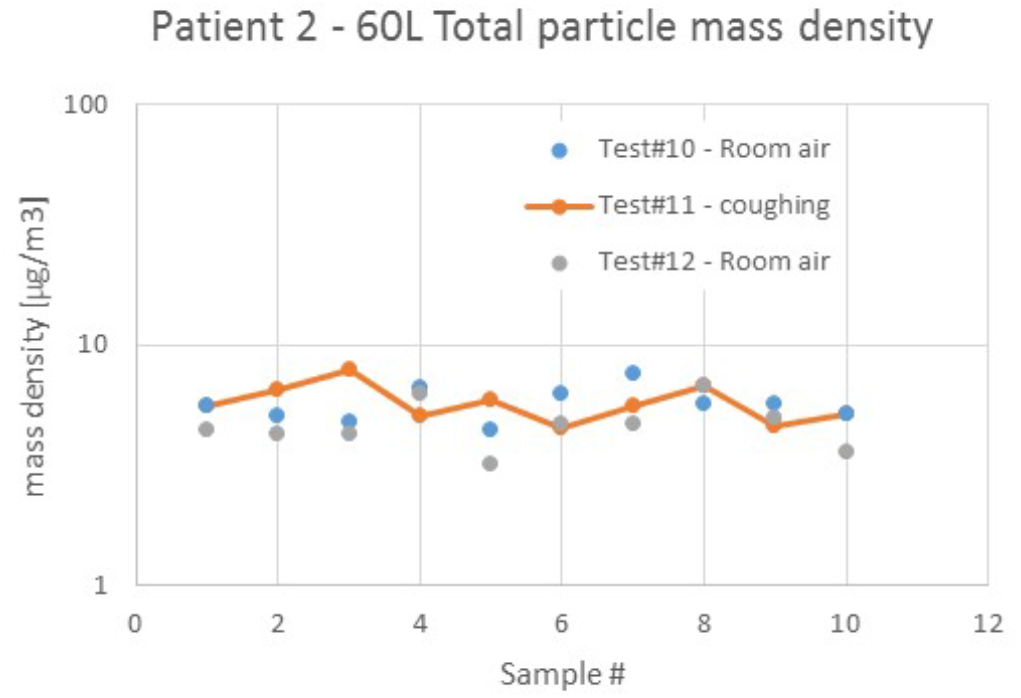
Particle mass concentration (PC) 60 L/min HHFNC and room air.

**Figure A4:**
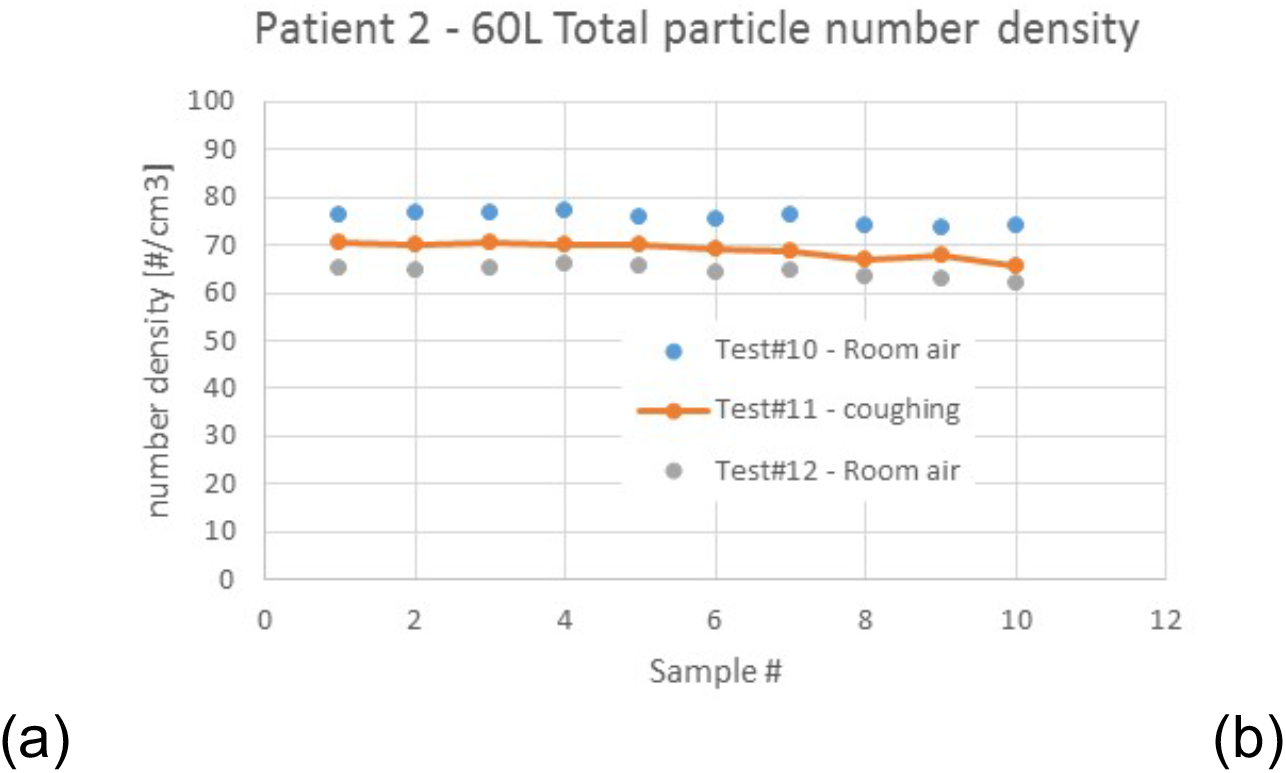
Particle number concentration (PC) 60 L/min HHFNC and room air.

**Figure A5:**
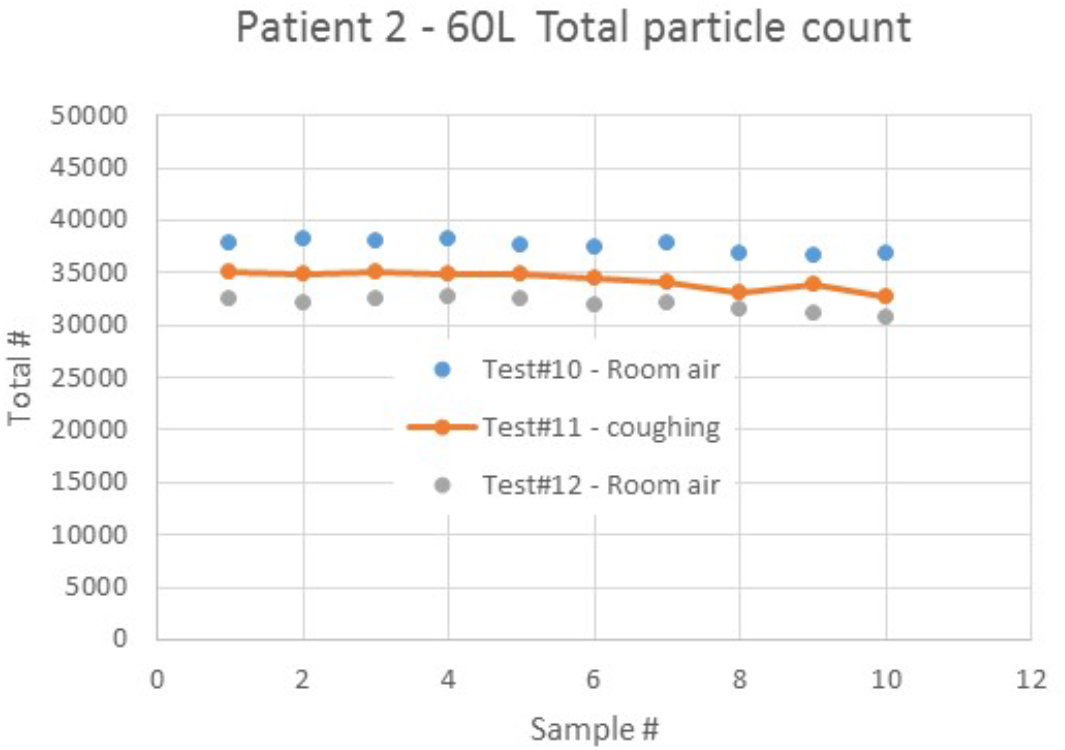
Total number of particles (PC) 60 L/min HHFNC and room air.

## Notes

### Competing Interest Statement

The authors have declared no competing interest.

### Clinical Trial

Not a clinical trial

### Funding Statement

Funding: None. This material is the result of work supported with resources and use of facilities at the Ann Arbor VA Medical Center.

